# Absolutely quantitated protein levels to reveal an ER/PR framework governing the full spectrum of breast cancer

**DOI:** 10.64898/2026.03.02.26347441

**Authors:** Guohua Yu, Junmei Hao, Jiandi Zhang, Fangrong Tang

**Author notes:** These authors contributed equally (Guohua Yu, Junmei Hao).

## Abstract

Cancer heterogeneity is traditionally attributed to multiple parallel signaling pathways. This belief is challenged here by proposing the ER/PR axis as the dominant pathway underlying the full spectrum of breast cancer. Absolutely quantitated ER, PR, Her2 and Ki67 protein levels were accumulated over 8 years from 1652 specimens collected non-selectively and measured with Quantitative Dot Blot (QDB) method over time. Cox analysis showed ER and Ki67 as independent adverse prognostic factors while PR was an independent favorable factor statistically. Their optimized stratification framework demonstrated that prognosis across all clinical subtypes was predominantly aligned along the ER/PR axis rather than being subtype-specific, including repeated identification of a subgroup with near-perfect 10-year survival probability from three independent cohorts to be proposed as the biological basis of the ultra-safe group in MINDACT trial. A parsimonious model is proposed where the ER/PR signaling hierarchy supersedes current prevailing clinical subtyping, with its balance essential for survival until ER levels become uncontrollable. This concept of pathway hierarchy may also exist in other major cancer types, and cannot be addressed without clinical epidemiology.

---

Cancer heterogeneity is widely accepted to arise from the activation of multiple signaling pathways across major types of cancer. For breast cancer, different signaling pathways are believed to underlie different clinical subtypes, with estrogen receptor (ER)/progesterone receptor (PR) signaling pathway supporting the luminal type, while Her2 signaling pathway dominating Her2-enriched (Her2) subtype. For triple negative breast cancer (TNBC), multiple signaling pathways co-exist to explain its diversified biology^1^.

In the past 8 years, we have accumulated 1,652 breast cancer specimens with matched clinical data, including survival status, from two hospitals in China non-selectively and continuously (**Supplemental Fig. 1**). Clinical subtypes were assigned by immunohistochemistry (IHC) to yield 10-year survival probability (10y SP) of 87% (n=312) for Luminal-A like (LumA); 80% (n=484) for Lumina-B like (LumB); 83% (n=291) for Her2; and 75% (n=168) for TNBC. 78 cases were classified as others due to unclassifiable or missing data (**Supplemental Fig. 2**). The protein biomarker levels of ER, PR, Ki67 and Her2 of these specimens were also measured absolutely and quantitatively using the Quantitative Dot Blot (QDB) method over time.

When cohort 1 (n=419) was used to demonstrate the reliability of QDB against IHC for stratification of LumA from LumB,^2–4^ the prognostic roles of ER, PR, Ki67 and Her2 were also investigated using Cox proportional hazards regression overall survival analysis treating absolutely quantitated protein levels as continuous variables with death as endpoint. As shown in **Supplemental table 1**, univariate Cox analysis identified both ER and Ki67 as adverse prognostic factors, with the hazard ratio (HR) increased by 1.61 per nmol/g for ER (95% CI: 1.24-2.10, p=0.0004) and 1.68 per 10 nmol/g for Ki67 (95% CI: 1.05-2.67, p=0.0302). Neither Her2 nor PR was observed as a prognostic factor with statistical significance. However, multivariable Cox analysis showed strengthened prognostic effects of ER, Ki67 and PR, with PR showing a nonsignificant favorable trend, to indicate an interdependent relationship (1.61 to 1.79 per nmol/g for ER, p=0.0000; 1.68 to 1.76 per 10 nmol/g for Ki67, p=0.0245; 0.9 to 0.82 per nmol/g for PR, p=0.1562). Thus, our data indicate ER, PR and Ki67 may be used to develop a potential classical interdependent prognostic signature. Notably, the proportional hazards (PH) hypothesis was violated for ER in both univariate and multivariate analysis, indicating its prognostic effect was not constant over time.

The unexpected adverse effect of ER was inconsistent with the existing literature, where ER was universally regarded as a favorable prognostic factor. However, ER was assessed in these studies either by IHC or ligand binding assay (LBA). IHC reflects the percentage of positively stained nuclei rather than total protein levels, while LBA measures the binding capacity of extracted cytosol. Our data, to the best of our knowledge, is the first to investigate its prognostic role by absolutely quantitated total protein levels in formalin fixed paraffin embedded (FFPE) tissues.

Optimized cutoffs for these biomarkers (1.01 nmol/g for ER, p<0.0001; 2.31 nmol/g for Ki67, p=0.00038; & 0.17 nmol/g for PR, p=0.0069) were identified using the “surv_cutpoint” function in R for maximum survival stratification **(Supplemental Fig. 3**). The PR cutoff was adjusted to its lower limit of quantitation (LLOQ) of 0.288 nmol/g for technical considerations.

For simplicity, we named those with ER levels <1.01 nmol/g as E_l_ vs ≥1.01 nmol/g as E_h_, Ki67 levels ≥2.31 nmol/g as K_h_ vs <2.31 nmol/g as K_l_, and PR levels ≥0.288 nmol/g as P_h_ vs <0.288 nmol/g as P_l_. Thus, the prognostic signature of ER<1.01 nmol/g, Ki67< 2.31 nmol/g and PR≥ 0.288 nmol/g should predict K_l_E_l_P_h_ subgroup with best prognosis of all cases to support findings from Cox analysis.

We identified 98 of 419 cases with this signature, including 5 deaths vs 82 deaths in the remaining 321 cases [10y SP: 94% (95% CI:89-99) vs 73% (95% CI: 68-78)] (**Supplemental Fig. 4a**). The K_l_E_l_P_h_ subgroup was identified again in another independent cohort (cohort 2) to include 92 cases with no deaths vs 5 deaths in the remaining 247 cases [5y SP:100% (95% CI: 100-100) vs 95% (95% CI: 88-100)] (**Supplemental Fig. 4b**). In another independent cohort (cohort 3), the KlElPh subgroup identified 69 cases with 1 death vs 85 deaths in the remaining 506 cases [10y SP: 99% (95%CI:96-100) vs 81% (95%CI: 77-85)] (**Supplemental Fig. 4c**).

Overall, we identified 259 cases as K_l_E_l_P_h_ group with 6 deaths and 10y SP at 97% (95%CI: 94%-100%) vs 78% (95%CI: 75%-81%) for the remaining 1074 cases with 172 deaths when all three cohorts combined (Fig.1). Examination of the 6 deaths in the K_l_E_l_P_h_ group showed that all cases had 2 or more lymph node metastases (n=2, 2, 4, 5, 7 and 11 respectively). In other words, if detected timely, this group could achieve 100% post-surgery survival in the real world.

**Fig 1:**
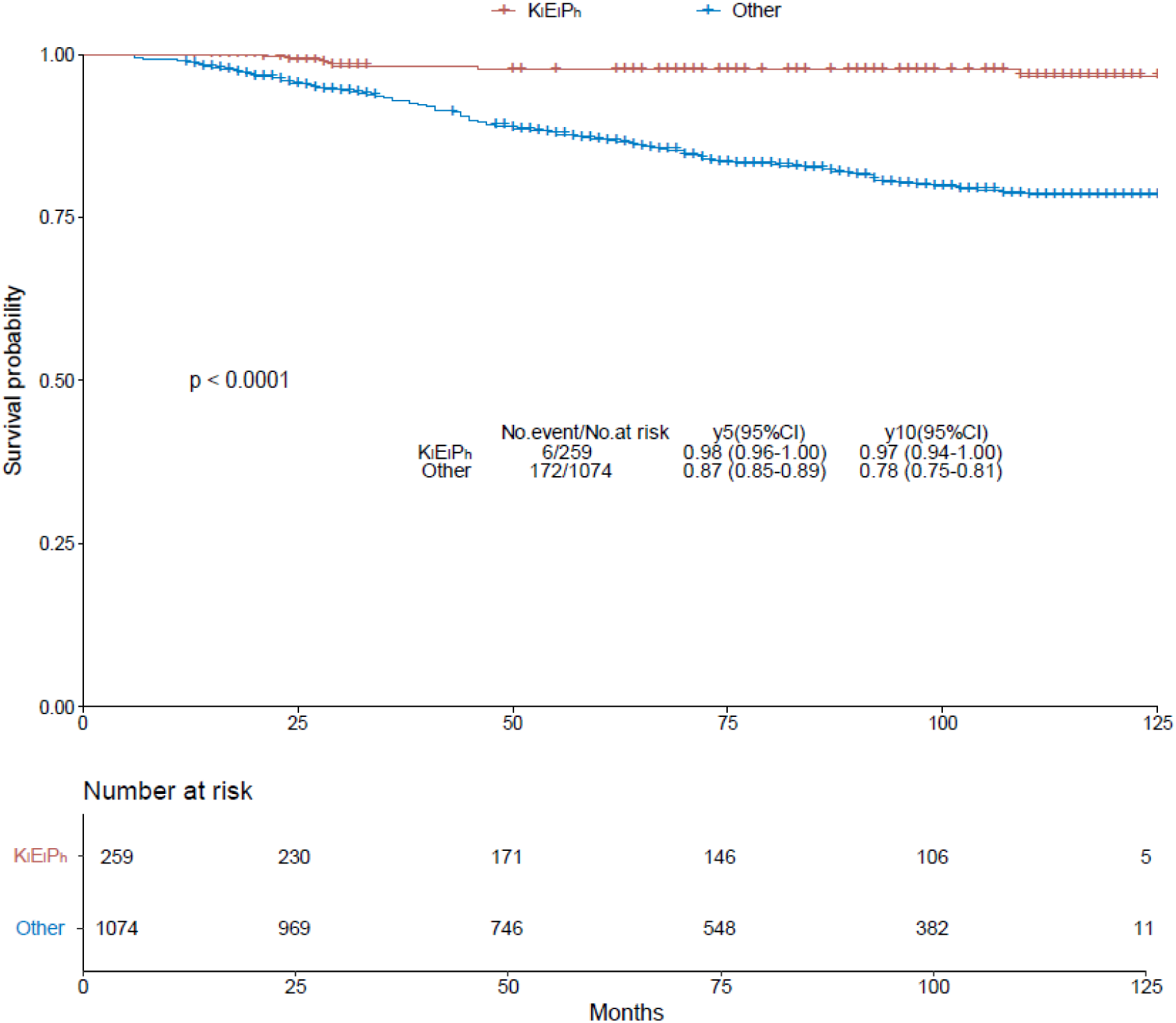
Kaplan Meier overall survival (OS) analysis of K_l_E_l_P_h_ vs the remaining cases in the combined cohorts of 1, 2 and 3. The cases were stratified with absolute quantitated ER, PR and Ki67 levels expressed as nmol/g, with ER <1.01 as E_l_ vs ≥1.01 as E_h_, Ki67 ≥2.31 as K_h_ vs <2.31 as K_l_, and PR ≥0.288 as P_h_ vs <0.288 as P_l_. Thus, **K_l_E_l_P_h_**with 10-year survival probability (10y SP) at 97% (95%CI: 94% −100%) is defined as cases of ER<1.01 & PR>0.288 & Ki67 <2.31 while 78% (95%CI: 75%-81%) for **Others** which include all remaining cases.

While unexpected, similar findings have already been described in the literature. In the recent MINDACT prospective trial of 6,693 patients, an ultra-safe group of 1000 cases was identified with an 8-year breast cancer-specific survival (BCSS) of 99.6% among patients with no more than 3 lymph node metastasis (N0 & N1).^5^ Notably, the MINDACT trial was tightly controlled with 93.7% BCSS even among high-risk groups. In our study, the overall 10y SP was only 81.7% as a real-world retrospective observation. When K_l_E_l_P_h_ subgroup was limited to N0 & N1, we identified 227 cases (2 deaths), accounting for 17% of total cases vs 14.9% of the ultra-safe group in the MINDACT trial. This similarity hints that we may look at the same group of patients.

The Cox analysis was repeated with the combined cohorts of 1333 cases (table 1). With significantly increased sample size, we observed again adverse effects of ER with HR at 1.25 per nmol/g (95%CI: 1.03-1.52, p=0.0247) and Ki67 with HR at 1.45 per 10 nmol/g (95%CI:1.06-1.98, p=0.0204). PR showed favorable role with HR at 0.82 per nmol/g (95%CI: 0.66-1.01, p=0.0672). The multivariate Cox analysis strengthened again the prognostic effects of all three biomarkers, with HR at 1.36 per nmol/g for ER (95% CI: 1.11-1.67, p=0.0024), 1.47 per 10 nmol/g for Ki67 (95%CI: 1.07-2.02, p=0.0162). Importantly, PR was demonstrated as a favorable factor with HR at 0.75 per nmol/g with statistical significance (95%CI: 0.59-0.96, p=0.0204). Her2 remains of no prognostic significance at this scale. Not surprisingly, for both ER and PR, the PH hypothesis was violated, suggesting their effects were time-dependent. Thus, our data clearly demonstrated ER, along with Ki67, were adverse prognostic factors while PR was a favorable factor for breast cancer.

**Table 1:** Cox proportional hazards regression overall survival analysis was performed with all cases (n=1333) using absolutely quantitated protein biomarker levels as continuous variables and death as end point. HR = hazard ratio; CI = confidence interval.

| Variables | Univariate |  |  | Multivariate |  |  |
| --- | --- | --- | --- | --- | --- | --- |
|  | HR | 95%CI | p-value | HR | 95%CI | p-value |
| ER (nmol/g) | 1.25 | 1.03-1.52 | 0.0247 | 1.36 | 1.12-1.67 | 0.0024 |
| Her2 (10 nmol/g) | 0.98 | 0.56-1.71 | 0.9445 | 0.96 | 0.55-1.68 | 0.8916 |
| Ki67 (10 nmol/g) | 1.45 | 1.06-1.98 | 0.0204 | 1.47 | 1.07-2.02 | 0.0162 |
| PR (nmol/g) | 0.82 | 0.66-1.01 | 0.0672 | 0.75 | 0.59-0.96 | 0.0204 |

When all cases were stratified by these three biomarkers, with E_h_K_l_ & E_h_K_h_ subgroup stratified by ER and Ki67 alone due to limited sample size, we observed the three biomarker combination offered better prognosis than prevailing clinical subtyping with p<0.0001 vs p=0.0048 (Fig. 2).

**Fig. 2:**
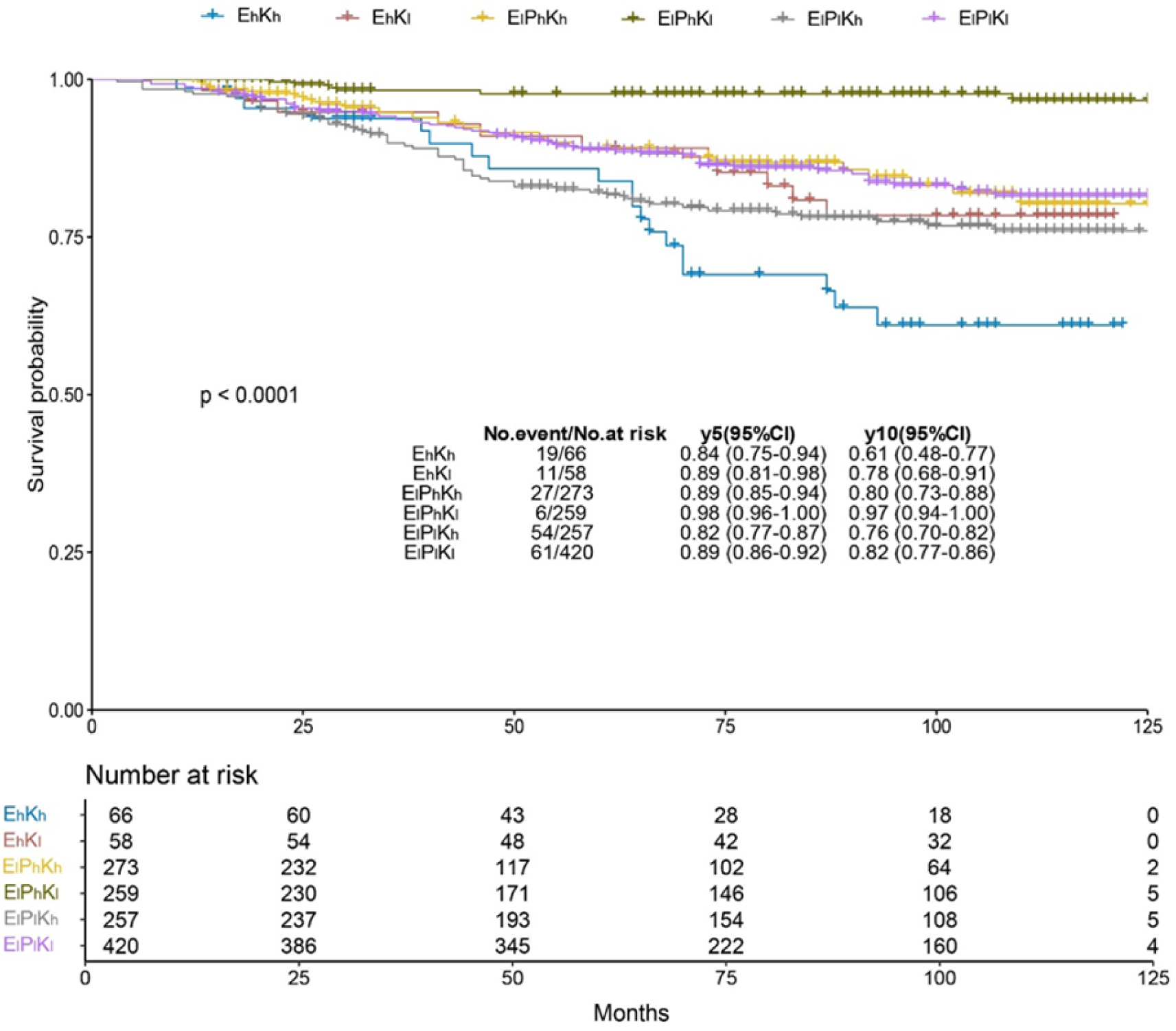
Kaplan-Meier overall survival analysis of cases based on absolute ER, PR and Ki67 protein levels. The values were expressed in nmol/g with ER <1.01 as E_l_ vs ≥1.01 as E_h_, Ki67 ≥2.31 as K_h_ vs <2.31 as K_l_, and PR ≥0.288 as P_h_ vs <0.288 as P_l_. We stratified Eh cases by Ki67 levels only due to limited sample size. Among 6 subgroups, K_l_E_l_P_h_ was observed with best prognosis (97%, 95%CI: 94%-100%) while E_h_K_h_ had the worst prognosis (61%, 95%CI: 48%-77%).

The patient characteristics of all subgroups were analyzed in **supplemental table 2**. We observed a tendency of more older patients (>50) in E_l_P_l_ subgroups and their almost exclusive presence in E_h_ group. Majority of patients were of pT1 and pT2, pN0 and N1, and being ductal invasive. As expected, the K_l_E_l_P_h_ group had the highest percentage of G_1_ patients while K_h_E_l_P_l_ had the highest percentage of G_3_ patients.

All four clinical subtypes were found in all subgroups. However, due to the inherent subjectivity and inconsistency of IHC, we arbitrarily treated subtypes accounting for <5% of total cases in a subgroup as misclassification. Accordingly, only K_l_E_l_P_l_ and K_h_E_l_P_h_ had all four subtypes with TNBC missing in K_l_E_l_P_h_; LumA missing in K_h_E_l_P_l_; TNBC missing in E_h_K_h_; and both TNBC and Her2 missing in E_h_K_l_ subgroup.

The presence of 135 LumA, 85 LumB, and 22 Her2 in K_l_E_l_P_h_ was unexpected, as it would challenge the clinical utility of the clinical subtype in ER/PR axis. The 22 Her2 cases were unlikely a misclassification with Her2 levels reached 13 nmol/g, 40 times over the validated cutoff based on IHC scores in some cases.^6,7^ For reference, there were also 3.3% Her2 positive cases in Ultra-safe group from MINDACT trial.^5^

The mortality ratio and 10-year restricted mean survival time (10y RMST) of individual subtype within K_l_E_l_P_h_ was evaluated to avoid potential bias associated with 10y SP. The overall mortality and 10y RMST were 2.3% (n=259) and 117.8 months to be near identical with 2.2% (n=135) and 118.3 months for LumA; 1.2% (n=85) and 118.6 months for LumB; and 0% (n=22) and 120 months for Her2.

In contrast, the mortality rate jumped significantly while 10y RMST dropped sharply at 14.5% (n=420) and 107.8 months overall for K_l_E_l_P_l_ subgroup vs 13.3% (n=90) and 110.6 months for LumA; 13.0% (n=131) and 109.1 months for LumB; 14.8% (n=108) and 107.3 months for Her2; and 20% (n=50) and 102.6 months for TNBC (**Fig. 3a**).

**Fig 3:**
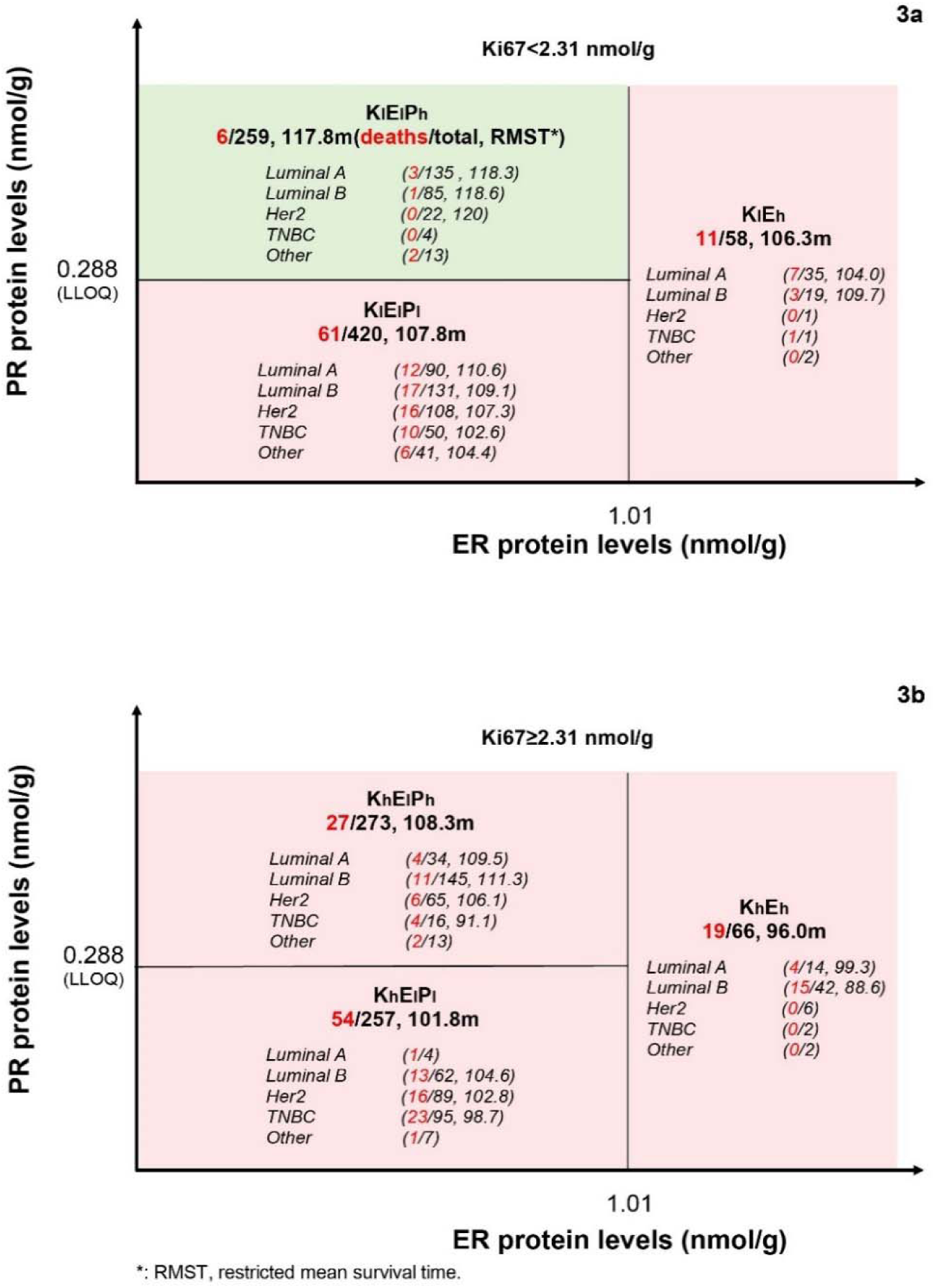
The restricted mean survival time (RMST) of individual subtypes within each subgroup defined by absolutely quantitated ER, PR and Ki67 protein levels. The mortality rate is indicated by the ratio of total deaths of total cases in a group or a subtype as specified in the figure. For any subtype accounting for less than 5% of total cases in a subgroup, only the mortality rate but not RMST is provided. **3a**, the mortality rate and RMST of individual subtypes within each subgroup of K_l_ group defined as cases with Ki67 levels <2.31 nmol/g. **3b**, the mortality rate and RMST of individual subtypes within each subgroup of Kh group defined as cases with Ki67 levels ≥2.31 nmol/g.

The largely subgroup-aligned rather than subtype-specific prognosis was best illustrated by LumA and LumB in these two subgroups. Their 10y RMST was nearly identical within the same subgroup, but drastically different between subgroups of same subtype (118.3 vs 118.6 in K_l_E_l_P_h_ vs 110.6 vs 109.1 in K_l_E_l_P_l_). Clearly, clinical subtype is overridden by the ER/PR framework in these subgroups.

This pattern was again confirmed in K_l_E_h_ subgroup, with 19.0% (n=58) and 106.3 months overall vs 20% (n=35) and 104 months for LumA; and 15.8% (n=19) with 109.7 months for LumB.

Each subgroup within the K_h_ group showed worse prognosis than its counterpart in K_l_ subgroups (**Fig. 3b**), and individual subtypes within each subgroup were again aligned largely with subgroup over subtype. For K_h_E_l_P_h_ subgroup, the mortality rate was 9.9% (n=273) with 10y RMST at 108.3 months overall vs 11.8% (n=34) and 109.5 months for LumA; 7.6% (n=145) and 111.3 months for LumB; 9.2% (n=65) and 106.1 months for Her2. The obvious exception of TNBC of 25% (n=16) with 91 months should not be over-interpreted, as high PR levels in this subgroup already contradict its clinical definition to be considered as misclassification of either LumA or LumB in this subgroup.

For K_h_E_l_P_l_ subgroup, it was 21% (n=257) and 101.8 months overall vs 21.0% (n=62) with 104.6 months for LumB, 18.0% (n=89) with 102.8 months for Her2 and 24.2% (n=95) with 98.7 months for TNBC. The K_h_E_h_ groups had the worst prognosis of 28.8% (n=66) and 96.0 months vs 28.6% (n=14) and 99.3 months for LumA, and 35.7% (n=42) and 88.6 months for LumB.

The largely subgroup-aligned rather than subtype-specific prognosis of the cases suggests the universal dominance of ER/PR framework over subtypes. Considering ER/PR levels are well-accepted surrogate biomarkers for ER/PR signaling pathway, our data suggested the ER/PR signaling pathway dominates the prognosis of the full spectrum of breast cancer. It inevitably places the ER/PR axis above the current prevailing clinical subtyping. We proposed that the prognosis of these subtypes is a reflection of their distribution alongside the ER/PR axis at the genetic level, the descriptive “phenotype” of ER/PR framework. Indeed, the intrinsic subtype concept was developed through microarray analysis. It is more descriptive in nature than being biology-driven^8^.

Due to the observational and retrospective nature, we lack sufficient treatment data to evaluate reliably its impact on the suggested ER/PR framework. Nonetheless, we analyzed the treatments each subtype received within K_l_E_l_P_h_ (**Supplemental table 3**) and K_l_E_l_P_l_ (**Supplemental table 4**) subgroups respectively. Among ∼40% cases with known treatments, we observed that the majority were treated with combined chemotherapy (CT) and endocrine therapy (ET) among LumA and LumB subtypes, and a fair number of LumA patients treated with CT alone in K_l_E_l_P_h_ subgroup. For Her2 subtype, at least 5 cases in K_l_E_l_P_h_ subgroup were not treated with Trastuzumab (TZD). Clearly, treatments alone were unable to explain this subgroup-aligned but not subtype-specific phenomenon.

The near perfect prognosis of K_l_E_l_P_h_ subgroup, regardless of the treatments, aligns well with MINDACT trial to support future de-escalation of treatments for these patients.^5^ It also supports strongly that ER/PR axis overrides subtyping in daily clinical practice. On the other hand, the slightly higher mortality rate and lower 10y RMST associated with both Her2 and TNBC over those of LumA and LumB may reflect the emergence of secondary signaling pathways like Her2 signaling pathway in K_l_E_l_P_l_ subgroups. Indeed, TZD treatment led to reduced mortality rate among Her2 subtype patients (1/12 vs 12/34). Thus, it raises a bigger question beyond current study: would effective TZD treatment achieve the prognosis of K_l_E_l_P_h_ or those of LumA and LumB of the same subgroup?

The drastic survival difference between the K_l_E_l_P_h_ and K_l_E_l_P_l_ group suggested that a tight balance of ER and PR expression levels is required to warrant patient survival if the tumors are detected timely. It thus aligned well with the putative role of PR at the molecular level.^9^ This balance is disrupted when ER protein levels become uncontrollable (>1.01 nmol/g), reflected by the worst prognosis of E_h_ group regardless of Ki67 overexpression. In fact, the K_h_E_h_ group was a previously unrecognized subgroup of worst prognosis. Active investigation is warranted in future.

It should be noted that current study provides a missing link required to unite the MINDACT trial and molecular PR rheostat model^5,9^ These two independent studies are disconnected across different realms of research, yet harmonize beautifully under the current ER/PR framework. It is a rare event for an observational study gaining support from both epidemiological and molecular signaling studies simultaneously to define the dominant role of ER/PR framework across the full spectrum of breast cancer.

Our results also challenge the prevailing view of ER/PR as good prognostic factors in breast cancer. To reconcile this clear discrepancy, we hypothesized a possibility that ER/PR balance may be reflected by the degree of luminal differentiation at the morphological level. IHC faithfully reflected the state of differentiation, but has until now been mistaken for total ER/PR protein levels in the tumor.

The revelation of ER/PR dominance in breast cancer would also challenge our perception of coexistence of multiple signaling pathways underlying the cancer heterogeneity to suggest the existence of a hierarchy of signaling pathways in breast cancer. Accordingly, restoring the collapsed dominant signaling pathway should be prioritized rather than spreading our efforts evenly among all signaling pathways for future clinical intervention. For example, prioritizing restoration of PR signaling pathway for K_l_E_l_P_l_ & K_h_E_l_P_l_ subgroups, which account for ∼50% of all cases when combined.^10,11^

If signaling hierarchy exists for breast cancer, the leading tumor for females accounting for 2 million new cases each year globally, could it also exist in other major cancers like lung, colorectal, gastric, and prostate? Obviously, this hypothesis can only be addressed through epidemiological studies, as re-creating the wide spectrum of cancer heterogeneity in real-world at the cellular, tissue and animal levels would be nearly impossible. We believe epidemiological studies addressing conceptual questions would become a new trend in future cancer research.

In summary, we demonstrated ER as an adverse prognostic factor by its total protein levels, and a dominant role of PR as a favorable regulator of breast cancer survival until ER levels become uncontrollable. By unifying all four clinical subtypes under a singular, quantitatively defined ER/PR/Ki67 framework, we demonstrate that breast cancer heterogeneity is not a collection of independent diseases, but a spectrum of failure under a dominant ER/PR framework for the whole spectrum of breast cancer. Thus, restoring PR signaling pathway would be the next logical step in future breast cancer research. More importantly, we postulated the likely presence of one dominant signaling pathway underlying other major types of cancer including lung, liver, colorectal and gastric to warrant further epidemiological investigations.

## Materials and Methods

### Patients

The inclusion criteria for this retrospective observational study were resection specimens from patients diagnosed with breast cancer with FFPE specimens archived in Yantai Yuhuangding Hospital and Yantai Affiliated Hospital of Binzhou Medical University at Yantai, China. The specimens had to have more than 50% tumor tissue based on H&E staining, and fixed in 10% neutral buffered formalin for 6-72 hours as recommended by national guidelines. All the procedures of the study were in accordance with the Declaration of Helsinki, and were approved by the medical ethics committee of Yuhuangding Hospital [2017]76 to guohua Yu and Yantai Affiliated Hospital of Binzhou Medical University (Approval #20191127001) to Junmei Hao; Follow-up data were obtained by cohorts, with the last follow-up at April 1, 2019 and March 31, 2020 for cohort 1; April 25, 2019 for cohort 2; February 25, 2022, April 3, 2022 and March 5, 2023 respectively for cohort 3.^2,3,6,7^ All clinical information was collected from patient records in the hospital with informed consent waived for archived specimens.

### General reagents

All of the chemicals were purchased from Sinopharm Chemicals (Beijing, P. R. China). Rabbit anti-ER (clone SP1, cat# ab13370) antibody was purchased from Abcam Inc. Anti-PR (clone 1E2, cat# 790-4296) rabbit monoclonal primary antibody was purchased from Roche Diagnostics GmbH. Rabbit anti-Her2 antibody (clone EP3, cat# ZA-0023) and mouse anti-Ki67 (clone MIB1, cat# ZM-0167) were purchased from ZSGB-BIO (www.zsbio.com). HRP labeled Donkey Anti-Rabbit IgG secondary antibody was purchased from Jackson Immunoresearch lab (West Grove, PA, USA). BCA protein quantification kit was purchased from Thermo Fisher Scientific Inc. (Carlsbad, CA, USA). QDB plates were provided by Quanticision Diagnostics, Inc. (RTP, USA).

### QDB assays

All QDB measurements were described in detail in other studies. In brief, 2X5 μm (or 2X15 μm if indicated in the reference) was used for protein extraction, with 0.5 μg/unit total protein lysates loaded to QDB plate alongside serially diluted protein standards and cell lysate controls with known protein biomarker levels. The plates were dried and then underwent a typical immunoblotting process including blotting, primary antibody and secondary antibody before the plates were incubated with ECL solution. The chemiluminescence signals were counted using a Tecan Infiniti 200pro Microplate reader with the option “plate with cover”.

### Statistical analysis

All statistical analyses were performed using R software (version 4.5.2). All statistical tests were two-sided, and a p value < 0.05 was considered statistically significant. Missing values in categorical variables were handled by introducing an additional category. Continuous variables were summarized as mean ± standard error of the mean (SEM).

Overall survival (OS) was defined as the time from surgery to death from any cause or the date of last follow-up. Patients who were alive at the last follow-up were censored. Patients lost to follow-up were excluded from the survival analyses.

Survival curves were estimated using the Kaplan–Meier method, and differences between groups were compared using the log-rank test. Univariate Cox proportional hazards models were used to estimate hazard ratios (HRs) and corresponding 95% confidence intervals (CIs) for OS. Multivariable Cox proportional hazards models were further constructed to evaluate the association between absolutely quantified protein biomarker levels and OS, with death as the endpoint. The proportional hazards assumption was assessed using Schoenfeld residuals.

Restricted mean survival time (RMST) was calculated for each group by integrating the Kaplan–Meier survival function up to a prespecified truncation time (τ = 120 months). RMST was defined as the area under the survival curve from time zero to τ. The variance of RMST was estimated using a Greenwood-type variance estimator, accounting for the cumulative uncertainty of the Kaplan–Meier estimator over the integration interval. Standard errors were derived accordingly, and two-sided 95% confidence intervals were constructed assuming asymptotic normality of the RMST estimator.

## Author contributions

Clinical samples & information, data analysis, manuscript editing: GY & JH; Conceptualization, drafting, editing & overall supervision: JZ; data analysis, editing and supervision: FT.

## Acknowledgements

We thank Jiahong Lyu & Zhenle Liu for technical assistance of current study

## Data statements

All data are accessible upon written request to

---

**Supplemental Fig. 1:**
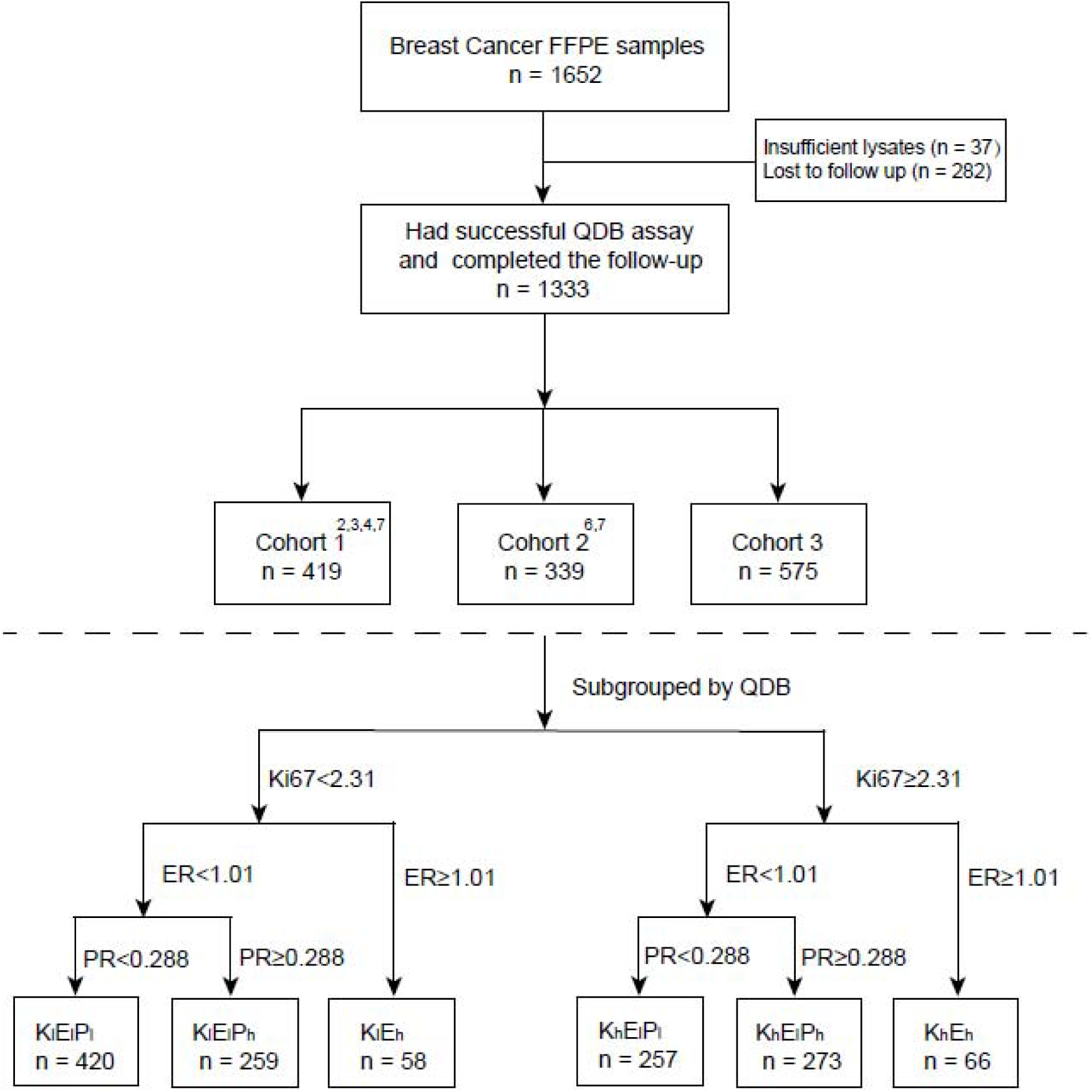
The flow chart of case selection for the study. Cohort 1 and 2 have been reported previously in other studies as indicated in the figure.

**Supplemental Fig. 2:**
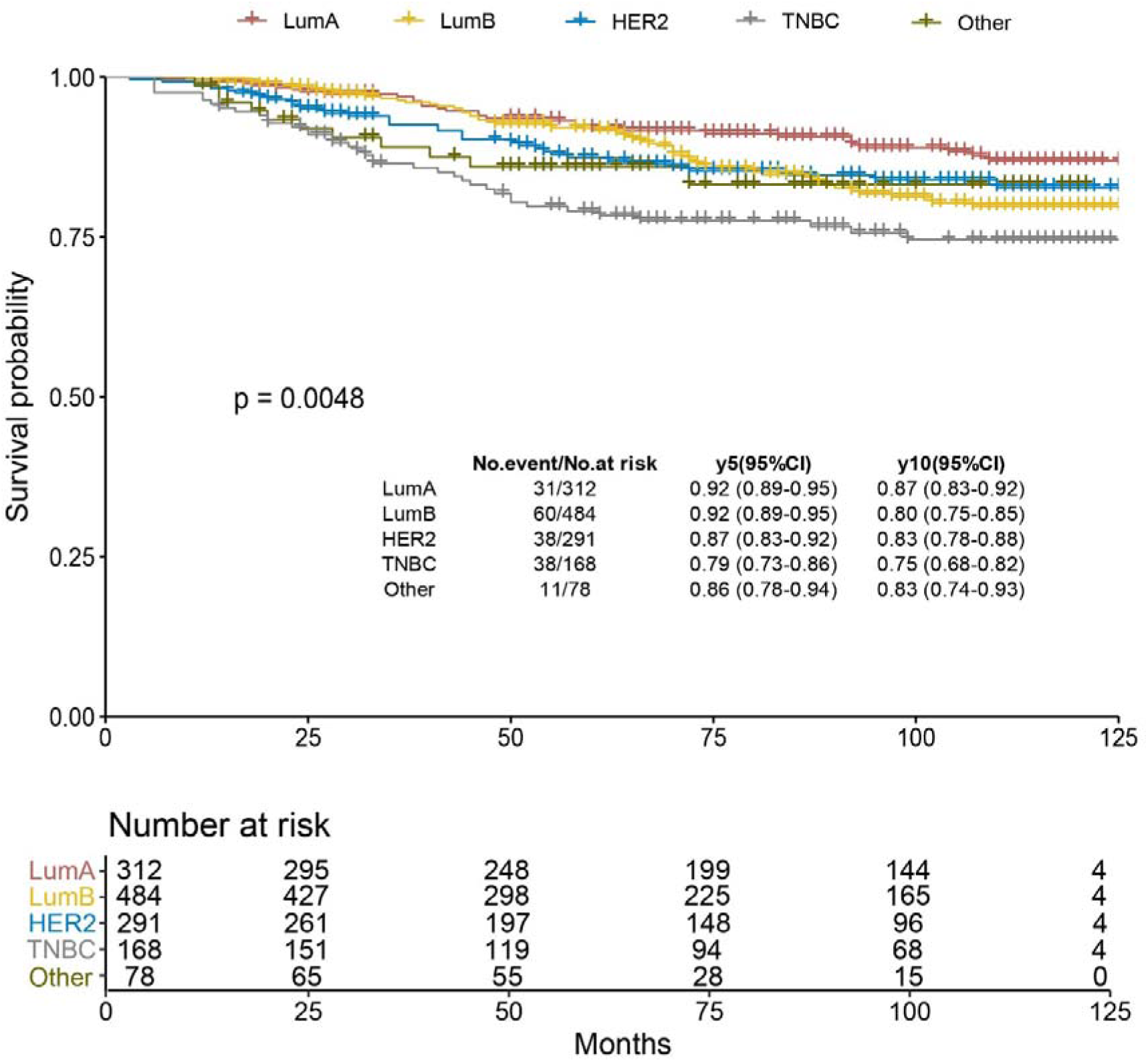
Kaplan-Meier analysis of overall survival (OS) of all cases stratified by clinical subtype based on the definition of 2013 St. Gallen consensus.^12^ In brief, Luminal A-like (LumA) is defined as ER and PR positive (ER+ & PR+), PR≥20%, Her2 negative (Her2-) and Ki67<20%; Luminal B (LumB) is defined as ER+, Her2-, PR<20% and/or Ki67≥20%; Her2-like (Her2) is defined as Her2+, and Triple Negative Breast Cancer (TNBC) is defined as ER-, PR-& Her2-. Cases not included in LumA, LumB, Her2 and TNBC, or cases with insufficient IHC information, were included in the Other category. For cases of Her2 2+, if there was no FISH result, we defined them as Her2+ or Her2-based on absolute Her2 protein levels using 0.26 nmol/g as cutoff defined in previous studies^6,7^.

**Supplemental Fig. 3:**
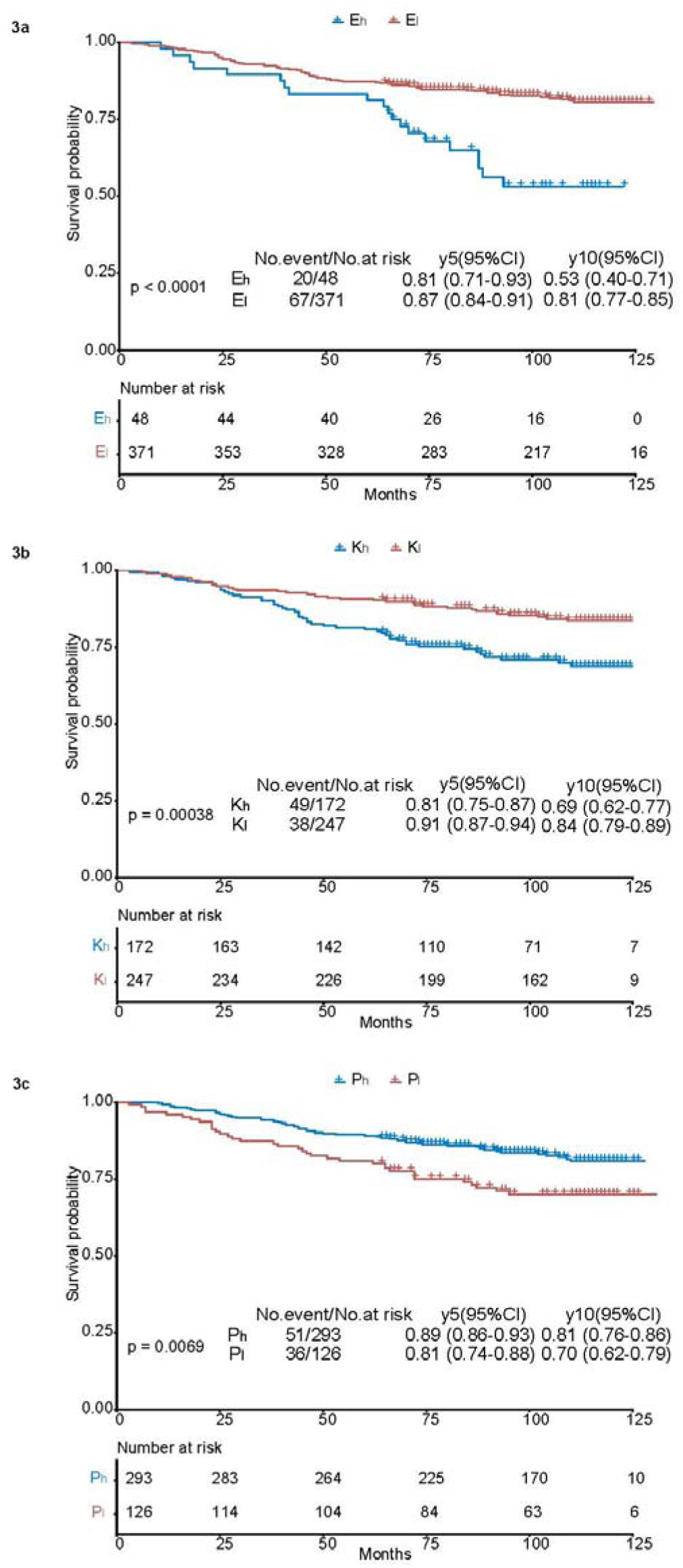
Developing optimized cutoffs of ER, PR, and Ki67 in cohort 1 (n=419) as training cohort 1. The optimized cutoff was identified using “surv_cutpoint” function of the “surviminer” R package based on absolutely quantitated protein biomarker levels and overall survival (OS). 3a, Optimized cutoff of 1.01 nmol/g was used to stratify patients into the E_h_ group for those with ER levels ≥1.01 nmol/g while those <1.01 nmol/g as E_l_ group, with p<0.0001; 3b, optimized cutoff of 2.31 nmol/g was used to stratify patients into K_h_ group with Ki67 levels ≥2.31 nmol/g while those <2.31 nmol/g as K_l_ group with p=0.00038; 3c, Optimized cutoff of 0.17 nmol/g was used to stratify patients into P_h_ group for those with PR levels ≥0.17 nmol/g while those <0.17 nmol/g as P_l_ group, with p=0.0069. For technical considerations, this cutoff was adjusted from 0.17 to 0.288 nmol/g, the lower limit of quantitation (LLOQ). LLOQ was defined as the lowest concentration level with both a CV < 25% and a relative error within ±25%, based on 40 calibration curves analyzed in triplicate.

**Supplemental Fig. 4:**
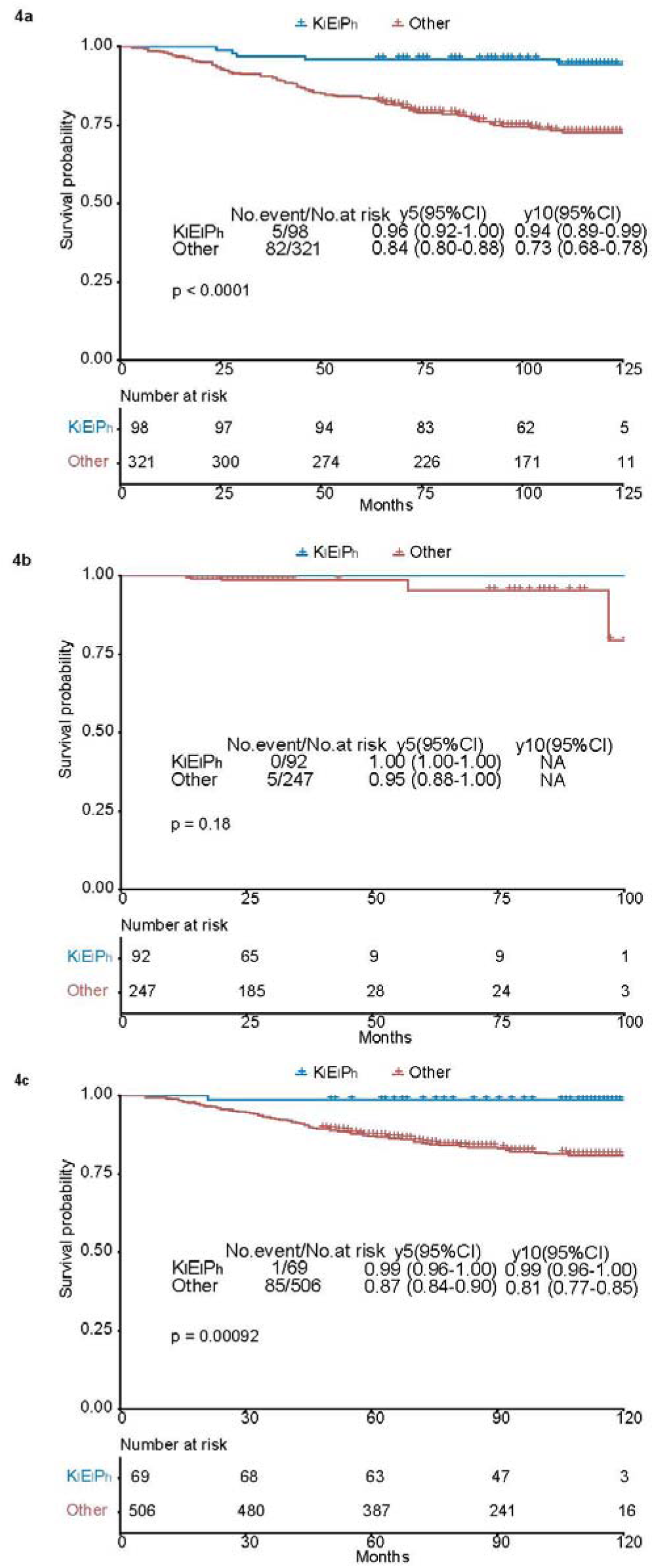
The Kaplan-Meier overall survival analysis of K_l_E_l_P_h_ subgroup in cohort 1, 2 and 3 respectively. The K_l_E_l_P_h_ subgroup was identified as cases with Ki67<2.31 nmol/g, ER<1.01 nmol/g, and PR≥0.288 nmol/g, and their overall survival (OS) was analyzed using Kaplan-Meier overall survival analysis vs the remaining cases in the same cohort. 4a, the Kaplan-Meier overall analysis of K_l_E_l_P_h_ vs the remaining cases in cohort 1 (n=419), with p<0.0001 using Log-rank test; 4b, the Kaplan-Meier overall analysis of K_l_E_l_P_h_ vs the remaining cases in cohort 2 (n=339), with p=0.18 using Log-rank test; 4c, the Kaplan-Meier overall analysis of K_l_E_l_P_h_ vs the remaining cases in cohort 3 (n=575), with p<0.00092 using Log-rank test;

**Supplemental Table 1:** Univariate and multivariate Cox proportional hazards overall survival regression analysis of cohort 1 (n=419) using absolutely quantitated ER, PR, Her2 and Ki67 protein levels as continuous variables with death as endpoint.

| Variables | Univariate |  |  | Multivariate |  |  |
| --- | --- | --- | --- | --- | --- | --- |
|  | HR | 95%CI | p-value | HR | 95%CI | p-value |
| ER (nmol/g) | 1.61 | 1.24-2.10 | 0.0004 | 1.79 | 1.36-2.36 | 0 |
| Her2 (10 nmol/g) | 1.43 | 0.77-2.68 | 0.2582 | 1.5 | 0.80-2.81 | 0.2033 |
| Ki67 (10 nmol/g) | 1.68 | 1.05-2.67 | 0.0302 | 1.76 | 1.08-2.87 | 0.0245 |
| PR (nmol/g) | 0.9 | 0.70-1.16 | 0.4189 | 0.82 | 0.62-1.08 | 0.1562 |

**Supplemental Table 2:**
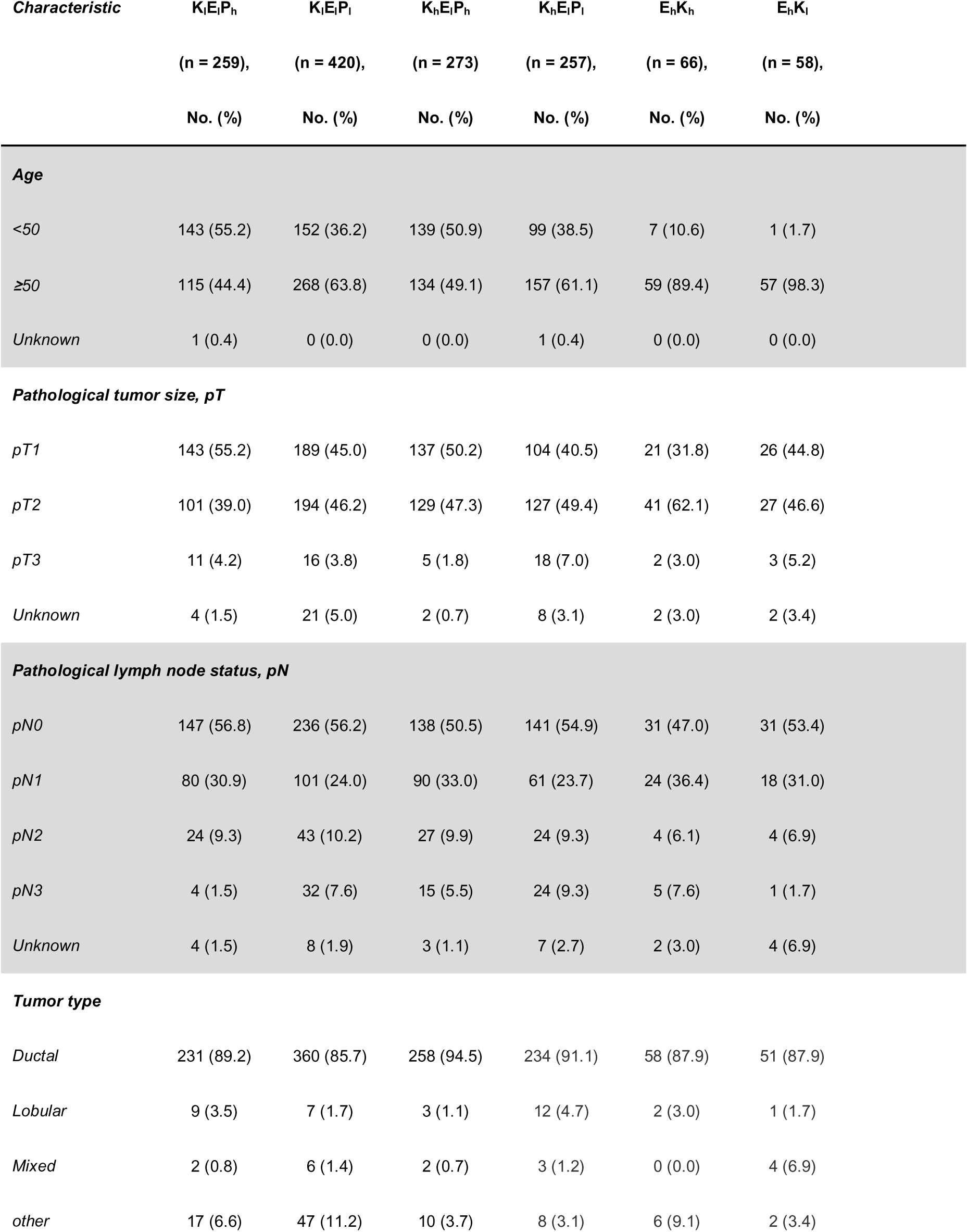
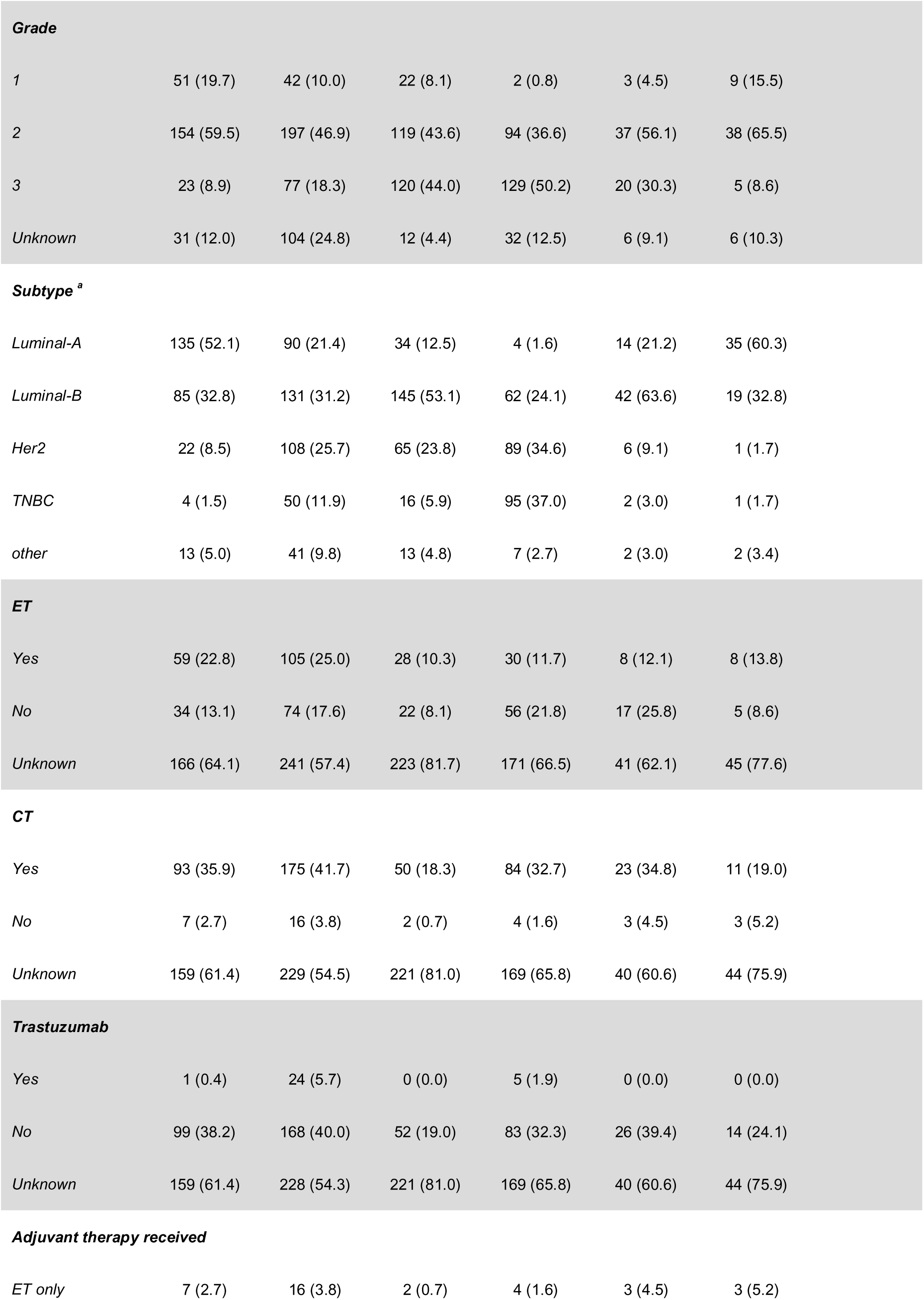
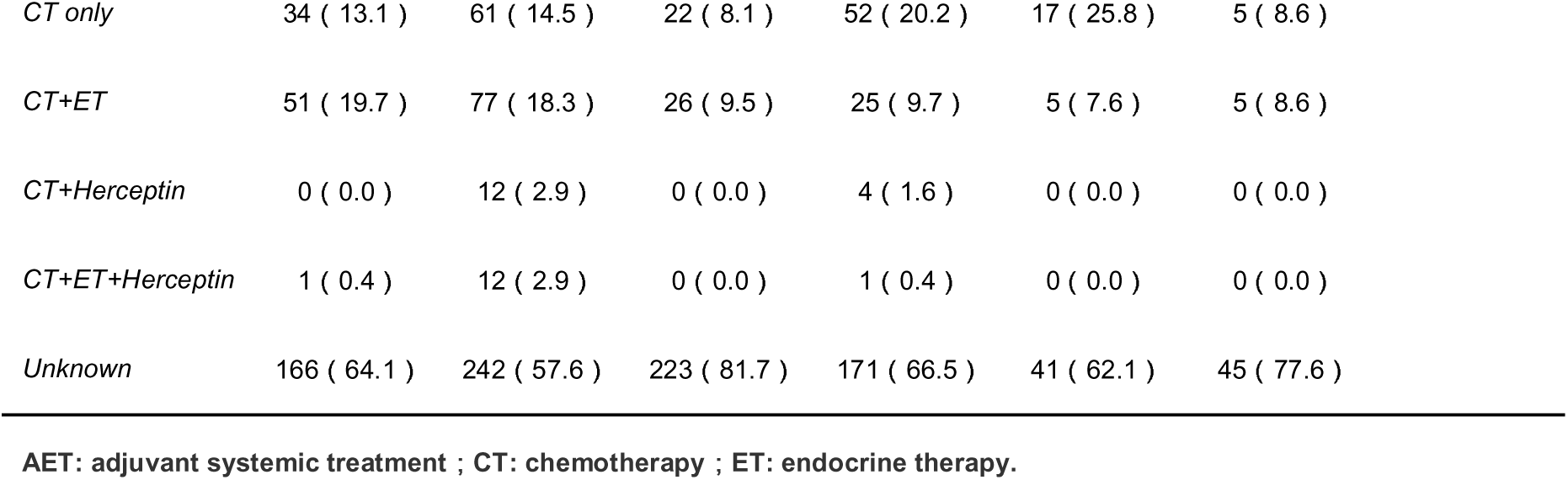
Patient characteristics by subgroup defined by absolutely quantitated ER, PR, Her2 and Ki67 protein levels.

**Supplemental table 3:** Summary of treatments received by individual subtype in K_l_E_l_P_h_ (3a) and K_l_E_l_P_l_ (3b) subgroup. ET: Endocrine therapy, CT: Chemotherapy. The number in () indicates number of deaths

|  | K<sub>i</sub>E<sub>i</sub>P<sub>h</sub> |  |  |  |  |
| --- | --- | --- | --- | --- | --- |
|  | LumA | LumB | Her2 | TNBC | other |
| ET only | 4(1) | 3 | 0 | 0 | 0 |
| CT only | 23 | 6 | 1 | 1 | 3(1) |
| CT+ET | 26(2) | 18(1) | 4 | 1 | 2(1) |
| CT+Herceptin | 0 | 0 | 0 | 0 | 0 |
| CT+ET+Herceptin | 1 | 0 | 0 | 0 | 0 |
| Unknown | 81 | 58 | 17 | 2 | 8 |

**3b**
|  | K<sub>i</sub>E<sub>i</sub>P<sub>i</sub> |  |  |  |  |
| --- | --- | --- | --- | --- | --- |
|  | LumA | LumB | Her2 | TNBC | other |
| ET only | 7(2) | 7 | 2 | 0 | 0 |
| CT only | 3 | 7 | 23(10) | 18(5) | 10(2) |
| CT+ET | 21(2) | 39(5) | 11(2) | 3(1) | 3 |
| CT+Herceptin | 0 | 0 | 6 | 2 | 4 |
| CT+ET+Herceptin | 0 | 2 | 6(1) | 0 | 4 |
| Unknown | 59(8) | 76(12) | 60(3) | 27(4) | 20(4) |

